# Patient and Hospital Characteristics Associated with Disparities in Acute Stroke Treatment in Community and Academic Hospitals

**DOI:** 10.1101/2023.09.25.23296119

**Authors:** Jerry Shen, Laura Corlin, Kathryn E. Coté, Megan E. Pudlo, Lester Y. Leung

## Abstract

**Importance:** Systemic biases and barriers may affect identification, emergency transportation, and care coordination for people with stroke. We assessed patient- and hospital-level factors for associations with pre-hospital and emergency department care processes. We compared trends for patients first presenting to an academic medical center versus community hospitals.

**Objective:** Assess whether patient and hospital characteristics were associated with differences in emergency medical services utilization, stroke code activation, and time to treatment for patients first hospitalized with stroke at academic medical center versus community hospitals. We hypothesized that disparities exist by patient characteristics (e.g., race and ethnicity, primary language) within each clinical setting, and that differences exist across the clinical settings.

**Design:** Retrospective cohort study using data from the electronic health record at an academic medical center (Tufts Medical Center).

**Setting:** Tertiary care referral center.

**Participants:** 542 patients aged ≥18 years hospitalized with stroke (96% with acute ischemic stroke) between 1/1/2018-12/31/2020, including patients who presented directly to the academic medical center and patients transferred from community hospitals.

**Main Outcomes:** Emergency medical services use, stroke code activation, door-to-computed tomography time, and door-to-needle time.

**Results:** Academic medical center non-Hispanic Asians (odds ratio (OR)=0.25; 95% confidence interval (CI)=0.13-0.47) and Hispanics (OR=0.19; 95% CI=0.05-0.72) and community hospital non-Hispanic Black/African-Americans (OR=0.17; 95% CI=0.05-0.62) were less likely to use emergency medical services compared to non-Hispanic whites. Patients with non-English primary language were less likely to use emergency medical services (OR=0.38; 95% CI=0.23-0.63) compared to English-speaking patients overall, in academic medical centers (OR=0.46; 95% CI=0.25-0.83) and community hospitals (OR=0.18; 95% CI=0.06-0.51). Community hospital Hispanics were less likely to have stroke code activation (OR=0.24; 95% CI=0.05-0.86) compared to non-Hispanic whites. Patients first presenting to a community hospital versus an academic medical center were less likely to have stroke code activation (OR=0.12; 95% CI=0.07-0.19), had shorter door-to-computed tomography time (31% shorter; 95% CI=15-43% shorter), and had longer door-to-needle time (29% longer; 95% CI=5-58% longer).

**Conclusions:** Patient-level factors and hospital setting were associated with differences in acute care suggesting opportunities for community outreach on emergency medical service use, interventions to alleviate language barriers, and approaches to address systemic racism affecting stroke care.

**Key Points:** *Question:* Are there differences in acute stroke care between patients first hospitalized at academic medical centers versus community hospitals?

*Findings:* In this retrospective cohort study (n=542), academic medical center patients identifying as non-Hispanic Asian or Hispanic, community hospital patients identifying as non-Hispanic Black/African-American, and patients with a non-English primary language were less likely to use emergency medical services. Community hospital patients identifying as Hispanic were less likely to have stroke code activation. Community hospital patients were less likely to have stroke code activation, had shorter door-to-computed tomography time, and had longer door-to-needle time.

*Meaning:* Targeted intervention and outreach are needed.

## Introduction

An essential aspect of stroke care is prompt transportation, identification, and coordination of care after symptom onset; however, systemic biases and issues such as cost of hospital transportation or delayed recognition of stroke symptoms can contribute to inequitable delays in treatment.^1^ Patient factors such as age, sex, and ethnicity have also been shown to correlate with disparities in treatment time across a variety of hospitals.^2^ These sources of disparity in treatment time can be assessed as targets for quality improvement in both community and academic hospitals so that more equitable treatment can be provided to all patients suffering from acute stroke.

Research utilizing quality improvement measures to reduce time to treatment has primarily focused on academic hospital settings, where the time from patient arrival to treatment is already shorter than average - in comparison, community hospitals (CHs) have been shown to have longer treatment times on average when compared to academic hospitals.^3^ National improvement initiatives involving a wide variety of hospitals have demonstrated effectiveness in larger academic hospitals but have less success in reducing times for other, smaller hospitals.^4^ We have not found much quantitative research assessing the different patient and hospital factors that are associated with differences in access to treatment or treatment time between community and academic hospitals.

Quantifying these differences and understanding the magnitude that patient and hospital factors play in contributing to these differences can bring to light areas of targeted improvement that CHs can address to provide more equitable care. At our tertiary care center (Tufts Medical Center, TMC), we see a diverse range of patients from the greater metropolitan area of Boston. We see both patients directly admitted to our academic medical center (AMC) and patients transferred after being initially seen at CHs for further management of acute stroke. We hypothesized that disparities exist by patient characteristics (e.g., race and ethnicity, primary language) within each clinical setting, and that there would be differences across the hospital settings.

## Methods

### Study Population

We conducted a retrospective cohort study using the Tufts Vascular Neurology Registry including patients age ≥18 years hospitalized at a comprehensive stroke/AMC with a primary diagnosis of acute ischemic stroke (AIS), transient ischemic attack (TIA), or stroke unspecified between 1/1/2018 and 12/31/2020 (eFigure 1). We included these three diagnoses reasoning that these patients would be treated during the early stages of acute stroke treatment like AIS. This cohort included two groups: patients who initially presented to the AMC for stroke care (n = 276) and patients transferred from CH emergency departments to TMC for further care (n = 266). This study was approved by the TMC Institutional Review Board (#12151).

### Predictors and Outcomes

Data extracted from the electronic health record included patient demographic characteristics (e.g., age at event [years], sex [female/male], race and ethnicity [Hispanic/non-Hispanic Asian/non-Hispanic Black or African-American/non-Hispanic White], primary language [English/other], insurance status [private, not private]), stroke risk factors (e.g., prior stroke, coronary artery disease, diabetes, hypertension, etc.), stroke severity and symptoms, and triage chief complaint. We also obtained participants’ ZIP Code level 2014-2018 GINI Index (a measure of neighborhood socioeconomic status assessing income inequality calculated using publicly available US Census Bureau data), and categorized patients as above or below the median GINI index in the United States as calculated in 2018 (0.414).^5,6^ ZIP Codes were determined based on area of residence documented during the time of admission. The date separating patients presenting before and during the COVID-19 pandemic was defined as the day that the World Health Organization defined COVID-19 a global pandemic – March 11, 2020.^7^ Additional details about variable definitions is included in online-only supplemental material.

There were four outcomes of interest: emergency medical services (EMS) utilization, stroke code activation, door-to-computed tomography (DTC) times, and door-to-needle (DTN) times. DTC time was the difference between the time of arrival to the emergency department of the initial admitting hospital and the time stamp of the first CT image obtained. DTN time was calculated as the time from arrival to the time of intravenous thrombolytic administration.

### Statistical Analysis

Pearson’s chi-squared test was used to assess whether participant characteristics varied by place of initial presentation (AMC or CH). We used logistic regression to assess associations with EMS utilization and stroke code activation, overall and stratified by place of initial hospitalization (AMC/CH). We used linear regression to assess associations with log-transformed DTC and DTN times. The primary models for EMS utilization were adjusted for place of initial presentation (unless stratified by this variable), age group (≥50/<50 years), sex (female/male), race and ethnicity (Hispanic/non-Hispanic Asian/non-Hispanic Black or African-American/non-Hispanic white), National Institute of Health Stroke Scale (NIHSS) score (≥5/<5), and presentation during the COVID-19 pandemic (during/before). Secondary models adjusted for the same variables but included primary language (English/other) instead of race/ethnicity.

The models for stroke code activation, DTC time, and DTN time were adjusted for all the same variables as the models for EMS utilization but also included EMS utilization (yes/no) and chief complaint (focal/non-focal). All analyses were conducted in R (version 4.1.3, Vienna, Austria).^8,9^

## Results

Baseline characteristics of the 542 included patients are shown in Table 1, stratified by location of initial hospital admission (51% first presented to the AMC). Overall, 65% used EMS, 62% had a stroke code called, the median DTN was 49 minutes, and the median DTC was 27 minutes. There were significant bivariate differences by location of initial hospital admission for EMS utilization, stroke codes, and mean log transformed DTC time but not mean log transformed DTN time. In bivariate analyses, patients who first presented to the AMC were significantly more likely to be ≥50 years, male, speak a non-English primary language, have a non-private insurance type, have a history of hyperlipidemia, have a focal chief complaint, and have a NIHSS score <5. There were also significant differences by race and ethnicity.

**Table 1.** Baseline characteristics of patients presenting first to an academic medical center (AMC) and patients presenting first to a community hospital (CH).

|  | **Total**<br>**(n = 542)**<br>**n (%)** | **AMC**<br>**(n = 276)**<br>**n (%)** | **CH**<br>**(n = 266)**<br>**n (%)** | **p value** |
| --- | --- | --- | --- | --- |
| **Method of Transportation, EMS** | 353 (65.1) | 158 (57.2) | 195 (73.3) | < 0.001 |
| **Stroke Code Called** | 336 (62.0) | 218 (79.0) | 118 (44.4) | < 0.001 |
| **Door to Computed Tomography Time**<br>**(Median Minutes, 25<sup>th</sup> Percentile, 75<sup>th</sup> Percentile)** | 27 (14, 58) | 31 (17, 63) | 24 (9, 53) | < 0.001 |
| **Door to Needle Time (Median**<br>**Minutes, 25<sup>th</sup> Percentile, 75<sup>th</sup> Percentile)** | 49 (33, 70) | 43 (29, 66) | 54 (39, 73) | 0.112 |
| ***Demographic Information*** |  |  |  |  |
| **Age < 50 (years)** | 54 (10.0) | 18 (6.5) | 36 (13.5) | 0.006 |
| **Female Sex** | 245 (45.2) | 113 (40.9) | 132 (49.6) | 0.042 |
| **Race and Ethnicity** |  |  |  | < 0.001 |
| Non-Hispanic White | 342 (64.0) | 137 (50.2) | 205 (78.5) |  |
| Non-Hispanic Asian | 111 (20.8) | 93 (34.1) | 18 (6.9) |  |
| Non-Hispanic Black/African-American | 48 (9.0) | 29 (10.6) | 19 (7.3) |  |
| Hispanic | 33 (6.2) | 14 (5.1) | 19 (7.3) |  |
| **Primary Language (Non-English)** | 122 (22.5) | 90 (32.6) | 32 (12.0) | < 0.001 |
| **Insurance Type, Not Private** | 284 (53.7) | 158 (59.2) | 126 (48.1) | 0.011 |
| **GINI Index of Residential ZIP Code**<br>**(2014-2018; ≤ 0.414, the US Median in 2018)** | 139 (26.1) | 63 (23.4) | 76 (28.8) | 0.158 |
| **During COVID-19 Pandemic** | 154 (28.4) | 80 (29.0) | 74 (27.8) | 0.764 |
| ***Prior Stroke History*** |  |  |  |  |
| **Prior Ischemic Stroke** | 65 (12.0) | 33 (12.0) | 32 (12.0) | 0.979 |
| **Prior Intracerebral Hemorrhage** | 7 (1.3) | 3 (1.1) | 4 (1.5) | 0.667 |
| **Prior Transient Ischemic Attack** | 41 (7.6) | 23 (8.3) | 18 (6.8) | 0.491 |
| **Prior Subarachnoid Hemorrhage** | 5 (0.9) | 3 (1.1) | 2 (0.8) | 0.683 |
| **Prior Subdural Hemorrhage** | 5 (0.9) | 3 (1.1) | 2 (0.8) | 0.683 |
| **Prior Intraventricular Hemorrhage** | 2 (0.4) | 2 (0.7) | 0 (0.0) | 0.164 |
| **Prior Cerebrovascular Accident, unspecified** | 24 (4.4) | 12 (4.3) | 12 (4.5) | 0.926 |
| ***Stroke Risk Factors*** |  |  |  |  |
| **Coronary Artery Disease/Myocardial Infarction** | 84 (15.5) | 39 (14.1) | 45 (16.9) | 0.370 |
| **Atrial Fibrillation** | 153 (28.2) | 70 (25.4) | 83 (31.2) | 0.131 |
| **Diabetes** | 164 (30.3) | 92 (33.3) | 72 (27.1) | 0.112 |
| **Hypertension** | 393 (72.5) | 200 (72.5) | 193 (72.6) | 0.981 |
| **Hyperlipidemia** | 266 (49.1) | 147 (53.3) | 119 (44.7) | 0.047 |
| **Obesity** | 40 (7.4) | 18 (6.5) | 22 (8.3) | 0.436 |
| ***Substance Use*** |  |  |  |  |
| **Current Tobacco Use** | 116 (21.4) | 56 (20.3) | 60 (22.6) | 0.520 |
| **Former Tobacco Use** | 121 (22.3) | 70 (25.4) | 51 (19.2) | 0.084 |
| **Alcohol Use** | 124 (22.9) | 56 (20.3) | 68 (25.6) | 0.144 |
| ***Initial Presentation*** |  |  |  |  |
| **Chief Concern, Focal** | 396 (73.1) | 212 (76.8) | 184 (69.2) | 0.045 |
| **Initial Systolic Blood Pressure >160** | 182 (33.6) | 95 (34.4) | 87 (32.7) | 0.673 |
| **Initial NIH Stroke Scale Score $\geq 5$** | 280 (52.4) | 127 (46.2) | 153 (59.1) | 0.003 |
| **Last Known Well Time to Emergency Department Arrival <4.5 hours** | 226 (42.0) | 111 (40.5) | 115 (43.6) | 0.474 |
| ***Stroke Diagnosis and Symptoms*** |  |  |  |  |
| **Diagnosis** |  |  |  | 0.130 |
| Acute Ischemic Stroke | 521 (96.1) | 261 (94.6) | 260 (97.7) |  |
| Transient Ischemic Attack | 19 (3.5) | 14 (5.1) | 5 (1.9) |  |
| Stroke, unspecified | 2 (0.4) | 1 (0.4) | 1 (0.4) |  |
| **Expressive Aphasia** | 156 (28.8) | 60 (21.7) | 96 (36.1) | < 0.001 |
| **Dysarthria** | 179 (33.0) | 101 (36.6) | 78 (29.3) | 0.072 |
| **Facial Weakness** | 189 (34.9) | 94 (34.1) | 95 (35.7) | 0.686 |
| **Arm Weakness** | 338 (62.4) | 157 (56.9) | 181 (68.0) | 0.007 |
| **Hemianopia** | 13 (2.4) | 5 (1.8) | 8 (3.0) | 0.363 |
| **Monocular Vision Loss** | 15 (2.8) | 8 (2.9) | 7 (2.6) | 0.850 |
| **Hemisensory Loss** | 69 (12.7) | 44 (15.9) | 25 (9.4) | 0.022 |
| **Hemiataxia** | 10 (1.8) | 5 (1.8) | 5 (1.9) | 0.953 |
| **Gait Ataxia** | 36 (6.6) | 23 (8.3) | 13 (4.9) | 0.107 |
| **Diplopia** | 11 (2.0) | 9 (3.3) | 2 (0.8) | 0.038 |

### Emergency Medical Services Utilization

Patient age, sex, and location of initial presentation (AMC versus CH) were not significantly associated with EMS utilization; however, NIHSS score, race/ethnicity, and primary language were (Table 2). Patients with NIHSS scores ≥5 were nearly 10 times as likely to use EMS, and this trend was similar in the results stratified by location of initial presentation. People who identified as a race and ethnicity other than non-Hispanic white were less likely to use EMS overall. Among patients presenting to the AMC, people identifying as non-Hispanic Asian (odds ratio (OR) = 0.25; 95% confidence interval = 0.13-0.47) and Hispanic (OR = 0.19; 95% CI = 0.05-0.72) were less likely to use EMS. Among patients presenting to CHs, people identifying as non-Hispanic Black/African American were less likely to use EMS (OR = 0.17; 95% CI = 0.05-0.62). Overall, preferring a language other than English was associated with a lower likelihood of using EMS (OR = 0.38; 95% CI = 0.23-0.63). This was somewhat more pronounced among patients presenting to CHs (OR = 0.18; 95% CI = 0.06-0.51) than to the AMC (OR = 0.46; 95% CI = 0.25-0.83). Having a stroke during the COVID-19 pandemic was associated with higher likelihood of using EMS for patients presenting to CHs but not to the AMC.

**Table 2.** Association of demographic and clinical factors with emergency medical services utilization of patients presenting first to an academic medical center (AMC) and patients presenting first to community hospitals (CH).

|  | **All Odds Ratio**<br>**(95 CI)** | **AMC Odds**<br>**Ratio (95 CI)** | **CH Odds Ratio**<br>**(95 CI)** |
| --- | --- | --- | --- |
| **Model 1** |  |  |  |
| **Age $\geq 50$** | 0.98 (0.47, 1.98) | 1.09 (0.32, 3.53) | 0.93 (0.34, 2.39) |
| **Female Sex** | 1.04 (0.67, 1.60) | 1.02 (0.56, 1.86) | 1.15 (0.59, 2.23) |
| **Race and Ethnicity (Non-Hispanic Asian)** | **0.33 (0.19, 0.58)** | **0.25 (0.13, 0.47)** | 1.06 (0.26, 5.55) |
| **Race and Ethnicity (Non-Hispanic Black/African-American)** | **0.40 (0.19, 0.85)** | 0.61 (0.23, 1.61) | **0.17 (0.05, 0.62)** |
| **Race and Ethnicity (Hispanic)** | **0.38 (0.16, 0.91)** | **0.19 (0.05, 0.72)** | 0.66 (0.20, 2.34) |
| **NIHSS<sup>a</sup> Score $\geq 5$** | **9.75 (6.26, 15.57)** | **9.22 (5.07, 17.54)** | **11.67 (5.84, 25.01)** |
| **During COVID-19 Pandemic** | 1.22 (0.77, 1.95) | 0.75 (0.40, 1.38) | **2.28 (1.08, 5.07)** |
| **CH Transfer** | 1.34 (0.85, 2.12) | - | - |
| **Model 2<sup>b</sup>** |  |  |  |
| **Non-English Primary Language** | **0.38 (0.23, 0.63)** | **0.46 (0.25, 0.83)** | **0.18 (0.06, 0.51)** |

<sup>a</sup> National Institutes of Health Stroke Scale
<sup>b</sup> Model 2 adjusted for the same variables as Model 1, except that language was used instead of race and ethnicity.

### Stroke Code Activation

Age, sex, primary language, NIHSS score, and stroke event during the COVID-19 pandemic were not associated with stroke code activation (Table 3). However, transferring from a CH was associated with lower likelihood of stroke code activation (OR = 0.12; 95% CI = 0.07-0.19).

**Table 3.**
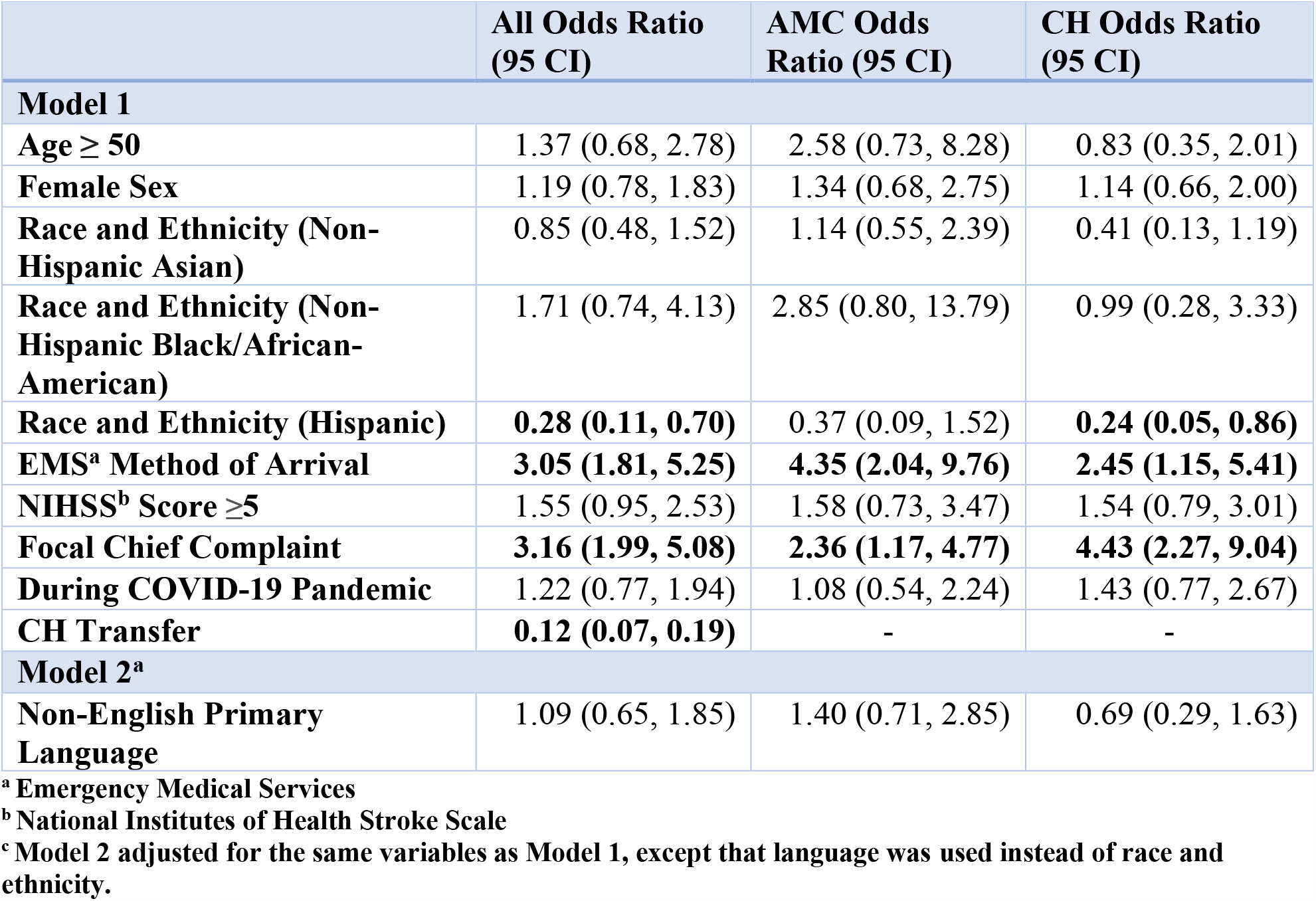
Association of demographic and clinical factors with stroke code activation of patients presenting first to an academic medical center (AMC) and patients presenting first to community hospitals (CH).

Using EMS and having a focal chief complaint were each associated with approximately three times the likelihood of stroke code activation. Similar trends for using EMS and having a focal chief complaint were observed in each of the AMC and CH groups. People identifying as Hispanic were less likely to have stroke code activation overall (OR = 0.28; 95% CI = 0.11-0.70) and if they first presented to a CH (OR = 0.24; 95% CI = 0.05-0.86).

### Door to CT Time

Initial presentation to CHs was associated with 31% shorter DTC times (OR = 0.69; 95% CI = 0.57-0.85; eTable 1). Overall and in models stratified by location of first presentation, EMS utilization, NIHSS score ≥5, and having a focal chief complaint were each associated with significantly shorter DTC time. For example, for patients first presenting to the AMC, EMS utilization was associated with 48% shorter DTC (95% CI = 33-59% shorter), NIHSS score ≥5 was associated with 24% shorter DTC (95% CI = 4-40% shorter), and having a focal chief complaint was associated with a 31% shorter DTC (95% CI = 11-47% shorter). For patients first presenting to a CH, EMS utilization was associated with 54% shorter DTC (95% CI = 32-69% shorter), NIHSS score ≥5 was associated with 47% shorter DTC (95% CI = 23-63% shorter), and having a focal chief complaint was associated with a 57% shorter DTC (95% CI = 40-70% shorter). Age, sex, race/ethnicity, and primary language were not associated with DTC overall, or in stratified analyses.

### Door to Needle Time

Overall, presenting to a CH first was associated with 29% longer DTN time (95% CI = 5-58% longer), having a non-English primary language was associated with 26% longer DTN time (95% CI = 0-59% longer overall), having a NIHSS score ≥5 was associated with a 34% shorter DTN time (95% CI = 14-49% shorter), and having a focal chief complaint was associated with 27% shorter DTN time (95% CI = 1-47% shorter; eTable 2). In analyses stratified by location of first presentation, only having a NIHSS score ≥5 was significantly associated with DTN time for patients first presenting to a CH (β = 58% shorter; 95% CI = 30-75% shorter). Age, sex, and race/ethnicity were not associated with DTN overall, or in stratified analyses.

## Discussion

We assessed disparities in the early, hyperacute care of patients with stroke. To our knowledge, this is one of the first studies comparing characteristics between stroke patients presenting to AMCs and CHs during this phase of care. Our results suggested differences by race and ethnicity for EMS utilization and stroke code activation and by primary language for EMS utilization and DTN time. Although neither age group nor sex were associated with any of the outcomes, clinical characteristics (e.g., NIHSS score and/or focal chief complaint) were associated with all the outcomes. Additionally, there were differences in EMS utilization, DTN time, and DTC time by location of initial presentation (an AMC in Chinatown of a major urban center or CHs in and near the urban center). There is a need for action to reduce the disparities observed.

Previous studies demonstrated disparities in stroke treatment according to race and ethnicity.^10,11,12^ Focusing on the hyperacute stage of care, our data are consistent with previous research demonstrating lower likelihood of EMS transport among patients identifying as a race and ethnicity other than non-Hispanic white.^12,13,14,15^ At least for our AMC results, the lower likelihood of EMS utilization among people who identify as non-Hispanic Asian may be due to the AMC’s location within Chinatown.^16^ However, we did not assess hospital proximity, and this may not explain other race and ethnicity trends we observed. Other possible explanations for lower utilization of EMS among people who identify as any race and ethnicity other than non-Hispanic white could include less recognition of stroke, distrust of EMS, and concern for cost.^17^

In addition to the racial and ethnic disparities observed for EMS utilization, we observed lower likelihood of stroke code activation among patients who identified as Hispanic, particularly among patients first presenting to CHs. As we did not observe differences by primary language for stroke code activation, the differences are unlikely to be explained by language barriers.

However, the trend is concerning and warrants further exploration. Conversely, and inconsistent with prior studies suggesting gender disparities in stroke code activation, we did not find differences by patient sex.^18^

Our observations of disparities by race and ethnicity may be an effect of systemic racism, suggesting an opportunity to implement interventions to address implicit bias and structural racism across the healthcare system. Previous work suggests that interventions could include community outreach and education to increase EMS utilization (e.g., emphasizing the health value and clarifying financial coverage options), awareness of stroke symptoms in minoritized populations, and expanded translation services to ensure timely and appropriate communication for care.^11,19^ Given the trends we observed for primary language, there could be language barriers affecting EMS utilization. These barriers may disproportionately affect minoritized patients.

EMS may need to provide higher quality in-ambulance translation services, as current methods to overcome language barriers have significant limitations.^20^ In our context, for example, outreach efforts around the AMC should focus on common languages spoken in the Boston Chinatown area. A hopeful message our results suggest is that once people are at the hospital, there were not racial and ethnic disparities at least in terms of time to imaging and treatment. This contrasts with previous studies demonstrating differences in treatment time associated with race and ethnicity.^12,21,22^

However, our study demonstrates that systemic disparities still exist with pre-hospital factors at the hospital level. Patients presenting to CHs first had less likelihood of stroke activation and longer DTN time (though they had shorter DTC time). Additionally, factors such as EMS utilization were more common during COVID-19 at CHs but not at AMCs. Both AMCs and CHs should carefully consider how interventions should be tailored for their local context.

This study has important strengths and limitations. First, the cohort was drawn from a large tertiary care referral center for a racially and socioeconomically diverse population of Massachusetts. Second, the cohort presenting to community hospitals drew from 45 unique hospitals in the Eastern Massachusetts region. Third, we have several different sets of outcomes utilizing a large set of variables describing individual-level demographic data. Fourth, we have three full years of data, including one year during COVID-19 which we were able to account for in our analysis. One limitation from this study is that data from community hospitals were drawn from transfer paperwork scanned into the EMR at Tufts, as we had limited access to patient data from the original hospitals. The provided transfer documentation may have been less thorough than that of patients presenting directly to Tufts. Another limitation is that this study draws only from stroke patients who were transferred from a community hospital and does not include those who remained at the original hospital to which they presented. Finally, we acknowledge that the discussion of race and ethnicity in this study is limited by the broad scope of the collective terms utilized when inputting demographic information in the electronic health record. We acknowledge that people can identify with more than one race and ethnicity.

## Conclusions

There are multiple intersecting disparities possible in pre-hospital and hospital-based hyperacute care for patients with stroke including race, ethnicity, and non-English primary language. These may represent opportunities for community outreach on EMS use, interventions to address systemic racism, and interventions to alleviate language barriers at both AMCs and CHs.

## Data Availability

All data produced in the present study are available upon reasonable request to the authors

## Acknowledgement

JS contributed data collection and analysis for this study, as well as the writing of the manuscript. LC contributed to the analysis as well as writing and editing assistance of the manuscript. KEC contributed data collection for this study and discussion of the use of GINI index in analysis. MEP contributed data collection for this study. LYL contributed guidance in data collection and analysis as well as writing and editing assistance of the manuscript. The authors have no conflicts of interest to disclose. Funding was provided for JS by the Committee of Medical Student Research at Tufts University School of Medicine via the Harold Williams, M.D. Summer Research Fellowship, which provides funding for medical students to conduct research during the summer between first and second year of medical school. Funding was also provided for LC by the Tufts University Data Intensive Studies Center. This information was previously partially presented in a poster presentation at the American Academic of Neurology Annual Meeting on April 25, 2023.

## Notes

### Competing Interest Statement

The authors have declared no competing interest.

### Funding Statement

This study was funded by the Committee of Medical Student Research at Tufts University School of Medicine via the Harold Williams, M.D. Summer Research Fellowship, which provides funding for medical students to conduct research during the summer between first and second year of medical school. Funding was also provided by the Tufts University Data Intensive Studies Center.

### Author Declarations

IRB of Tufts Medical Center gave ethical approval for this work (#12151).

