## Supplementary material for "Patient and Hospital Characteristics Associated with Disparities in Acute Stroke Treatment in Community and Academic Hospitals": Online Supplement

### Online-Only Supplement

eFigure 1. CONSORT Flow Diagram

eTable 1. Association of demographic and clinical factors with Door-to-Computed Tomography time of patients presenting first to an academic medical center (AMC) and patients presenting first to community hospitals (CH).

eTable 2. Association of demographic and clinical factors with Door-to-Needle time of patients presenting first to an academic medical center (AMC) and patients presenting first to community hospitals (CH).

Variable Definitions

### eFigure 1. CONSORT Flow Diagram

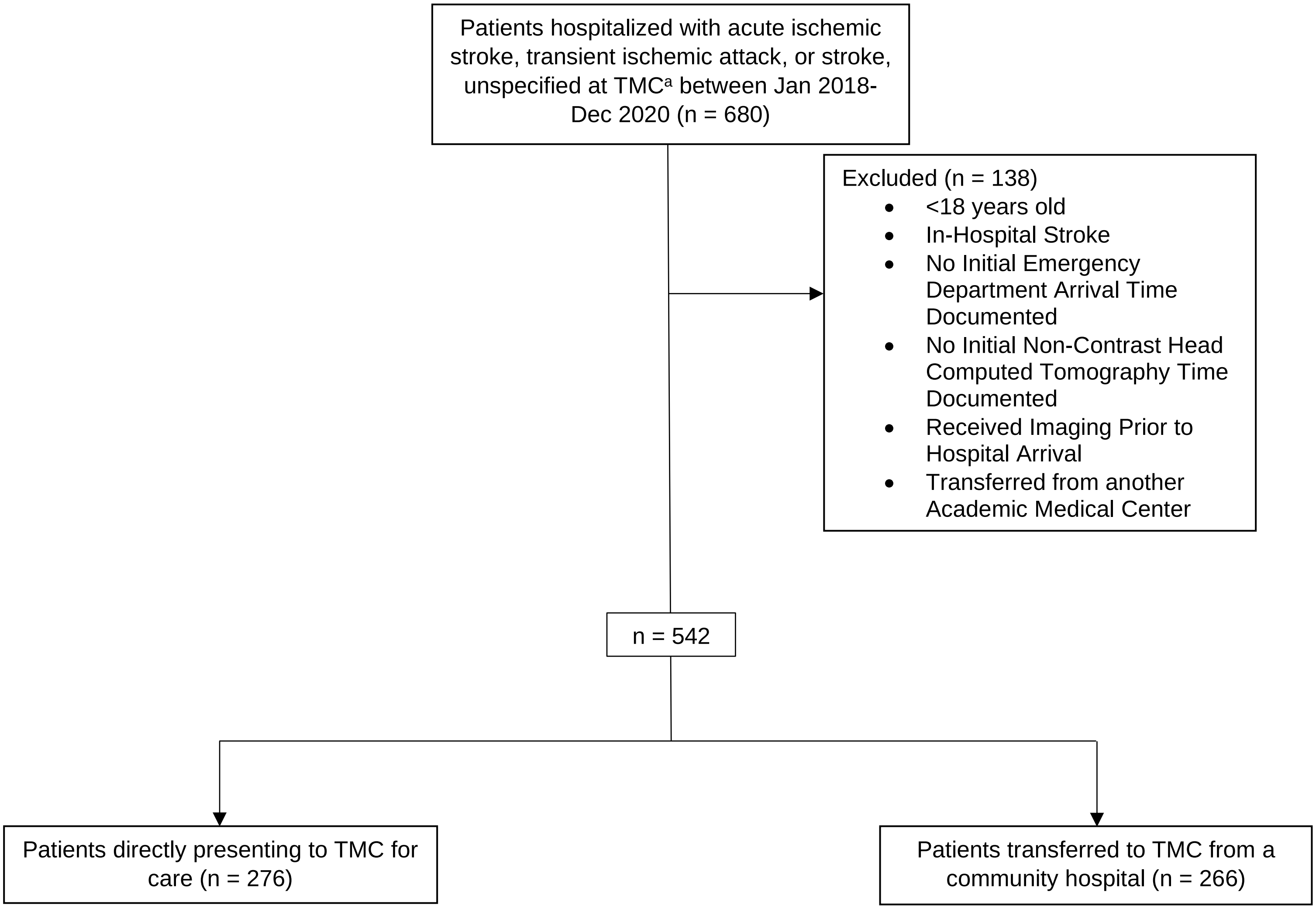


[a]Tufts Medical Center

**eTable 1. Association of demographic and clinical factors with Door-to-Computed Tomography time of patients presenting first to an academic medical center (AMC) and patients presenting first to community hospitals (CH).**

| | | All<br>Exponentiated Coefficient (95% Confidence Interval) | AMC<br>Exponentiated Coefficient (95% Confidence Interval) | CH<br>Exponentiated Coefficient (95% Confidence Interval) |
|---|---|---|---|---|
| **Model 1** | | | | |
| **Age ≥ 50** | | 1.05 (0.76, 1.44) | 1.07 (0.69, 1.65) | 1.08 (0.67, 1.74) |
| **Female Sex** | | 1.10 (0.92, 1.33) | 1.11 (0.89, 1.37) | 1.09 (0.81, 1.49) |
| **Race and Ethnicity** | | | | |
| | Non-Hispanic Asian | 1.00 (0.78, 1.28) | 1.15 (0.90, 1.46) | 0.81 (0.44, 1.49) |
| | Non-Hispanic Black/African-American | 1.07 (0.76, 1.50) | 1.01 (0.71, 1.44) | 1.20 (0.63, 2.29) |
| | Hispanic | 1.35 (0.91, 2.00) | 1.60 (0.97, 2.64) | 1.13 (0.62, 2.06) |
| **EMS[a] Transportation** | | 0.50 (0.40, 0.62) | 0.52 (0.41, 0.67) | 0.46 (0.31, 0.68) |
| **NIHSS[b] Score ≥ 5** | | 0.62 (0.51, 0.77) | 0.76 (0.60, 0.96) | 0.53 (0.37, 0.77) |
| **Focal Chief Complaint** | | 0.52 (0.42, 0.65) | 0.69 (0.53, 0.89) | 0.43 (0.30, 0.60) |
| **During COVID-19** | | 1.02 (0.83, 1.24) | 1.07 (0.85, 1.35) | 1.03 (0.73, 1.45) |
| **CH Transfer** | | 0.69 (0.57, 0.85) | - | - |
| **Model 2[c]** | | | | |
| **Non-English Primary Language** | | 0.92 (0.73, 1.15) | 0.94 (0.75, 1.18) | 0.98 (0.61, 1.57) |

[a] Emergency Medical Services
[b] National Institutes of Health Stroke Scale
[c] Model 2 adjusted for the same variables as Model 1, except that language was used instead of race and ethnicity

**eTable 2. Association of demographic and clinical factors with Door-to-Needle time of patients presenting first to an academic medical center (AMC) and patients presenting first to community hospitals (CH).**

| | | All<br>Exponentiated Coefficient (95% Confidence Interval | AMC<br>Exponentiated Coefficient (95% Confidence Interval) | CH<br>Exponentiated Coefficient (95% Confidence Interval) |
|---|---|---|---|---|
| **Model 1** | | | | |
| **Age ≥50** | | 1.15 (0.79, 1.66) | 1.07 (0.66, 1.72) | 1.41 (0.70, 2.85) |
| **Female Sex** | | 1.20 (0.99, 1.46) | 1.26 (0.96, 1.65) | 1.10 (0.81, 1.50) |
| **Race and Ethnicity** | | | | |
| | Non-Hispanic Asian | 1.08 (0.82, 1.42) | 1.02 (0.74, 1.41) | 1.26 (0.63, 2.52) |
| | Non-Hispanic Black/African-American | 0.83 (0.54, 1.27) | 0.77 (0.48, 1.23) | 1.17 (0.31, 4.50) |
| | Hispanic | 1.38 (0.95, 2.01) | 1.19 (0.68, 2.09) | 1.57 (0.92, 2.68) |
| **EMS[a] Transportation** | | 0.86 (0.62, 1.18) | 0.72 (0.48, 1.08) | 1.30 (0.72, 2.34) |
| **NIHSS[b] Score ≥ 5** | | 0.66 (0.51, 0.86) | 0.80 (0.59, 1.09) | 0.42 (0.25, 0.70) |
| **Focal Chief Complaint** | | 0.73 (0.53, 0.99) | 0.71 (0.46, 1.10) | 0.81 (0.48, 1.38) |
| **During COVID-19** | | 1.02 (0.82, 1.25) | 1.07 (0.80, 1.42) | 0.97 (0.69, 1.35) |
| **CH Transfer** | | 1.29 (1.05, 1.58) | - | - |
| **Model 2[c]** | | | | |
| **Non-English Primary Language** | | 1.26 (1.00, 1.59) | 1.20 (0.90, 1.60) | 1.41 (0.92, 2.16) |

[a] Emergency Medical Services
[b] National Institutes of Health Stroke Scale
[c] Model 2 adjusted for the same variables as Model 1, except that language was used instead of race and ethnicity.

### Variable Definitions

Age, sex, race, ethnicity, and primary language were determined from the patient demographics page in each patient's chart. During analysis, all languages other than English were grouped into the category "Other Language." Insurance type was obtained from the patient demographics page and separated into two categories: "Private" and "Not Private." If the patient had any sort of private insurance, they were categorized as "Private." If the patient had public health insurance or were non-insured, they were categorized as "Not Private." The 2014-2018 GINI index is a measure of neighborhood socioeconomic status assessing income inequality calculated using publicly available US Census Bureau data. GINI Index was separated into two categories: above or below the median GINI index in the United States as calculated in 2018 (0.414).[23] All methods of transportation that did not involve emergency medical services (EMS) were categorized as "Walk-in" including private vehicles or ride share. Past medical and social history known upon patient arrival was obtained from the initial encounter, either at a community hospital or at Tufts Medical Center. The Chief Complaint was obtained from patient charts, determined by initial documentation by the triage nurse. The Chief Complaint was divided into two categories, "Focal" or "Non-Focal" based on the chief concern. Focal chief complaints were defined as those chief complaints describing focal neurological deficits, such as aphasia, vision loss, hemineglect, unilateral weakness (face, arm, and/or leg), unilateral sensory loss (face, arm, and/or leg), and ataxia. Chief complaints such as "stroke symptoms" or "stroke code" were also included in the focal category. Initial systolic blood pressure and National Institute of Health Stroke Scale (NIHSS) score were obtained from initial encounter notes. The last known well time to first emergency department arrival was calculated as the difference between the door time and the last known well time as documented in the patient's initial encounter note. The presence of stroke symptoms as described by the patient or their family were elicited from the initial encounter documentation.

Whether a stroke code was called or not was determined from the initial encounter documentation. If documentation was not clearly indicated, the patient was documented as not having a stroke code called. Whether the patient had pre-hospital notification (e.g., "Stroke Alert") by EMS was also determined from the initial encounter documentation. If documentation was not clearly indicated, the patient was documented as not having pre-hospital notification. Patients who arrived via walk-in methods of transportation were excluded from analysis relating to pre-hospital notification. The door time was defined as the time that the patient arrived at the first site of care, as indicated in the initial encounter note. Door-to-Computed Tomography (CT) time was calculated as the difference between the door time and non-contrast head CT time (the time stamp present in the CT imaging). Door-to-Needle time was calculated as the difference between the door time and the time of needle puncture for intravenous thrombolytic.
